# PanelCAT: an Open-Source Comparative Analysis Tool for Next-Generation Sequencing Panel Target Regions

**DOI:** 10.1101/2023.06.19.23291594

**Authors:** André Oszwald, Lucia Zisser, Eva Compérat, Leonhard Müllauer

## Abstract

Multi-gene next-generation sequencing (NGS) panels have become a routine diagnostic method in the contemporary practice of personalised medicine. To avoid inadequate test choice or interpretation, a detailed understanding of the precise panel target regions is required. However, the necessary bioinformatic expertise is not always available, and publicly accessible and easily interpretable analyses of target regions are scarce. To address this critical knowledge gap, we present the Panel Comparative Analysis Tool (PanelCAT) an open-source application to analyze, visualize and compare NGS panel DNA target regions. PanelCat uses RefSeq, ClinVar and COSMIC cancer mutation census databases to quantify the exon and mutation coverage of target regions and provides interactive graphical representations and search functions to inspect the results. We demonstrate the utility of PanelCAT by analyzing two large NGS panels (Illumina TSO500 and Qiagen pan-cancer panel) to validate the advertized target genes, quantify targeted exons and mutations, and identify differences between panels. PanelCat will enable institutions and researchers to catalogue and visualize NGS panel target regions independent of the manufacturer, promote transparency of panel limitations, and share this information with employees and requisitioners.

## Introduction

Precision oncology routinely involves next-generation sequencing (NGS) of tumor DNA to identify therapeutically actionable targets or diagnostically relevant mutations that critically direct patient management^1^. Most multi-gene sequencing panels do not cover entire genes, but only variable portions of genes that are considered most relevant, i.e., predominantly protein-coding sequences and tumor mutational hotspots. For this reason, both the choice of an adequate test and its interpretation, especially regarding the certainty of negative findings, crucially depend on detailed knowledge of the portions of genes and genetic alterations that may be assessed by a panel.

Target regions of commercial NGS panels are typically specified in a panel-specific BED file by a list of chromosome numbers, start and stop coordinates^2^. Although this information is an essential part of the test documentation, it is not useful to understand panel target regions in detail without further analysis for several reasons: it does not inform on the non-targeted portions of genes without comparison to a reference genome; the provided information on target genes, transcripts and exons is not updated alongside the transcript databases (e.g., RefSeq^3^); the target regions must be systematically compared to mutation/variant databases in order to determine pathogenic mutations that can be detected; lastly, genomic positions with known high rates of erroneous variant calls are often masked during secondary analysis, but these positions are defined in separate files.

Consequently, the lack of detailed publicly available data on precise panel targets, and the barriers to generate it due to the required bioinformatic expertise, portend the risk of inadequate test choice and test misinterpretation. To reduce this risk, we developed the “Panel Comparative Analysis Tool” (PanelCAT), an application that allows to analyze, visualize and compare DNA target regions of NGS panels within a user-friendly interface, and provides a platform to clearly communicate this information to others. We demonstrate the use of this tool by analyzing two large multi-gene NGS panels, the Illumina TrueSight Oncology 500 (TSO) and Qiagen Pan-Cancer (QPC) Panel, in order to provide a more detailed documentation of their targeted genes, exons, known pathogenic mutations, and differences between the panels, than has been available to date.

## Materials and Methods

### PanelCat code

PanelCat code was created, and all analyses were performed, in R^4^ v4.3.0 within RStudio v2023.03.0. Analysis of genomic ranges (target regions and variant coordinates) were performed using the GenomicFeatures^5^ package. Graphs were drawn using ggplot2^6^ and plotly^7^ packages. A browser-based implementation of the script was created using ShinyR^8^. PanelCat is provided under the open-source license AGPLv3 and the source code, and R session information is available at https://github.com/aoszwald/panelcat. The basic procedure of the panel analysis is outlined below.

PanelCat accepts target region files as input (containing columns for chromosome, start, and end position of target regions), and optionally the mask region file (also containing chromosome, start and end coordinates). The application first determines the intersection between the panel target regions and RefSeq^3^ exon coordinates in order to systematically identify target genes, and subsequently all exon ranges of target genes. The exon ranges of each targeted gene are then intersected with the panel target regions to quantify the targeted portion of protein coding bases per gene. Targeted mutations are then identified by intersection of the panel target regions with the coordinates of mutations in the ClinVar^9^ and COSMIC^10^ databases. Optionally, the mask file is incorporated in the analysis to identify and determine the portion of masked bases and mutations. The summarized output data are combined into lists of items and saved as R data objects. Panels that were previously analyzed and saved in this form are pre-loaded the next time the application is started. The panel output files can also be used for analysis outside of PanelCat; within R, the panel data and listed sub-items can be accessed via the “$” operator. In the course of this study, individual data were accessed in this way and further processed in R independently of the PanelCat functions to answer specific questions, including the cumulative percentage of non-targeted mutations, and discrepancies between the advertized gene list and the confirmed gene list.

### Data sources

BED files indicating target regions of NGS panels and corresponding mask files, including Illumina TSO500 and Qiagen Pan-Cancer Panels, were obtained from the customer support of the manufacturers, or obtained in the course of using a product. The TSO500 mask file was provided by Illumina. These files are not provided as part of PanelCat.

The following databases are not provided as part of the software download, and need to be either downloaded manually (COSMIC) or automatically by PanelCat (ClinVar and ClinVar assembly report, RefSeq):

ClinVar data (last accessed 25.05.2023) was obtained from https://ftp.ncbi.nlm.nih.gov/pub/clinvar/vcf_GRCh37/weekly/clinvar.vcf.gz. The COSMIC cancer mutation census data (v98, last accessed 23.05.2023) was obtained from https://cancer.sanger.ac.uk/cosmic/download. RefSeq data (last accessed 25.05.2023) was obtained from https://ftp.ncbi.nlm.nih.gov/refseq/H_sapiens/annotation/GRCh37_latest/refseq_identifiers/GRCh37_latest_genomic.gff.gz, and the GrCh37. The NCBI assembly report was obtained (last accessed 25.05.2023) from https://ftp.ncbi.nlm.nih.gov/genomes/refseq/vertebrate_mammalian/Homo_sapiens/annotation_releases/105.20220307/GCF_000001405.25_GRCh37.p13/GCF_000001405.25_GRCh37.p13_assembly_report.txt.

## Results

### General function of PanelCat

PanelCat (https://github.com/aoszwald/panelcat and https://aoszwald.shinyapps.io/panelcat/) provides functions to automatically distill descriptive information from panel target region files and public databases, and to display this data to facilitate evaluation and comparison of panels. To analyze a panel, PanelCat is provided with the target region file (typically with a .bed file suffix, but others may be acceptable), and optionally a mask file (indicating regions where variant calls are unreliable and will be filtered out). The application then determines the overlap between target regions and protein-coding bases per gene in RefSeq^3^, known pathogenic and likely pathogenic mutations in ClinVar^9^ and tier 1-3 oncogenic mutations in COSMIC^10^ cancer mutation census (CMC) databases. The output data is saved in a compact form that can be used in PanelCat or explored independently in R Statistics^4^.

The application provides several visualization options to analyze and compare panels, briefly outlined here. The first option is a scatter plot, where users can contrast target coverage metrics of RefSeq, ClinVar and COSMIC databases from all analyzed panels. This function can be used to compare between panels, e.g., to determine differences in target gene coverage (Fig. 1A), or within panels, e.g., to evaluate the relationship between coverage of protein-coding bases and pathogenic mutations. The second option is a horizontal column plot showing per-gene coverage, and the masked portion, of protein-coding bases, ClinVar variants and COSMIC mutations, whereby users can select multiple panels for comparison (Fig. 1B). The third visualization is a column plot optimized to search for and display one or multiple specific genes of interest across several (or all) analyzed panels, with indication of a panel that targets all searched genes (Fig. 1C). The fourth visualization is a column plot of the estimated frequency of COSMIC CMC mutations that are not targeted by the panels, both in target genes only and in all genes, with and without considering variant masking (Fig. 1D). Lastly, PanelCat provides a graph to compare base coverage of individual exons per transcript between two panels in paired violin plots (Fig. 1E).

**Figure 1:**
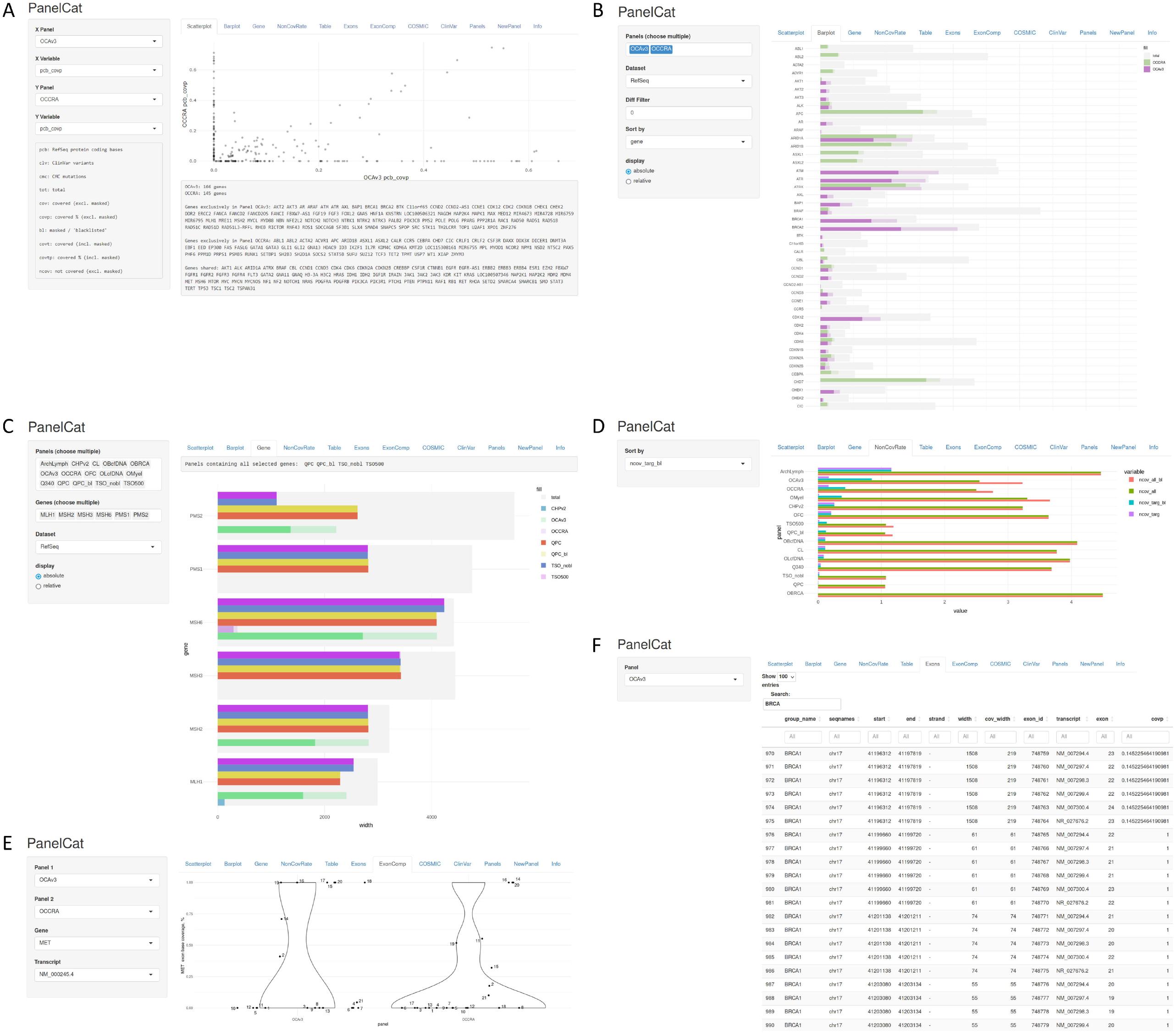
PanelCat user interface and visualisation options. Users can select tabs with several graphs to investigate panel DNA target region coverage. Options include an X/Y scatterplot to compare any coverage metric from any previous analysed panel (A), barplots to view or compare absolute or relative RefSeq exon base/ClinVar variant/COSMIC mutation coverage per gene (B), a search function to compare specific genes of interest across multiple panels (C), a graph to visualise the estimated average rate of non-targeted COSMIC CMC tier 1-3 mutations (D), violin plots to compare coverage per exon for a transcript of interest between two panels (E), multiple table views to inspect Exons (F), ClinVar variants or COSMIC CMC mutations.

Besides graphs, PanelCat provides table views to compare raw numerical data, or investigate the coverage of specific exons and mutations. In the first option, users can inspect the PanelCat output data (representing the raw data from which the graphs are generated), whereby multiple panels and metrics can be simultaneously displayed and searched for specific genes. The second option displays the coverage of each exon of every transcript of all target genes of a panel (Fig. 1F). Similarly, the third and fourth options provide for each panel a complete list of targeted ClinVar variants and COSMIC mutations, including the ability to display masked variants. The exon, ClinVar and COSMIC tables can be searched/filtered for each column independently, e.g., for specific genes, transcripts, exons, coding or amino acid changes, or genome coordinates.

When run locally, PanelCat automatically obtains the current ClinVar (released weekly) and RefSeq databases upon first use. These, along with all previously processed panels, can be updated in a single step; previous versions of databases and panel analyses are stored for later reference and documentation. The COSMIC CMC database (updated every several months) requires manual download and replacement of the local file.

In summary, PanelCat offers multiple useful and intuitive functions to substantially improve the transparency and accessibility of NGS panel target region documentation. The newly developed tool was next used to analyze two very large panels currently used in both clinical and research settings; the Illumina TrueSight Oncology 500 (TSO) and the Qiagen Pan-Cancer (QPC) panel. Although the target regions can be requested as part of the panel documentation, the explicit coverage of genes and mutations are not provided. However, informed clinical use requires detailed information, so we used PanelCat to characterize their target regions in detail and explore their subtle differences.

### Mutation coverage in Qiagen PanCancer panel is similar or greater than TSO500, despite lower exon coverage

In their respective product documentation, the TSO and QPC panels advertise the same 523 gene targets for analysis of small variants (e.g, SNV, insertions and deletions). We first compared the advertized genes to the target genes identified using PanelCat. The QPC target regions overlapped with exons of 603 genes, including all advertized genes. By contrast, the TSO target regions overlapped with exons of 625 genes, but these included only 521 of the 523 advertised targets. The two remaining target genes (HLA-B and HLA-C) do not overlap with the TSO target regions; accordingly, we did not find any variant calls in HLA-B or HLA-C in a representative set of 400 samples analyzed with the TSO panel (including unfiltered variant call files in 10 samples). Alterations in HLA-I genes have been postulated to promote tumor evasion of immune surveillance, e.g., by restricting neoantigen presentation^11,12^, although no guidelines recommendations to test HLA genes exist momentarily.

We next identified the targeted exon-coding bases of each gene. We first searched for genes with the greatest coverage and found that in the TSO, 20 genes had exon coverage > 95% (including NAB2, TERC, CD74, TFE3, KIF5B, EML4, EWSR1, FLI1, ETV1, ETV5, PAX3, all over 99%), whereas in the QPC, it was only six (TERC, ZRSR2, ATR, POLD1, KMT2B, RECQL4). We found a strong direct correlation between the base coverage of TSO and QPC panels (Pearson’s r = 0.81, p < 2e-16), and no significant difference in mean exon base coverage per gene (TSO 50.3% vs. QPC 48.4%, p = 0.23) (Fig 2A). We identified target genes where relative coverage was considerably greater in the TSO than in the QPC, including NTRK2, ETV1, AKT3, ERG, and PAX7, but only few genes with greater coverage in the QPC panel, notably PMS2, TERT, HLA-B and HLA-C (Fig. 2B). Importantly, four targeted genes (HLA-A, KMT2B, KMT2C, KMT2D) showed total masking of all target regions in the TSO panel (but not in the QPC, which does not use a mask file). Accordingly, we did not find any variants calls in these genes in a representative set of 400 samples analyzed with the TSO panel.

**Figure 2:**
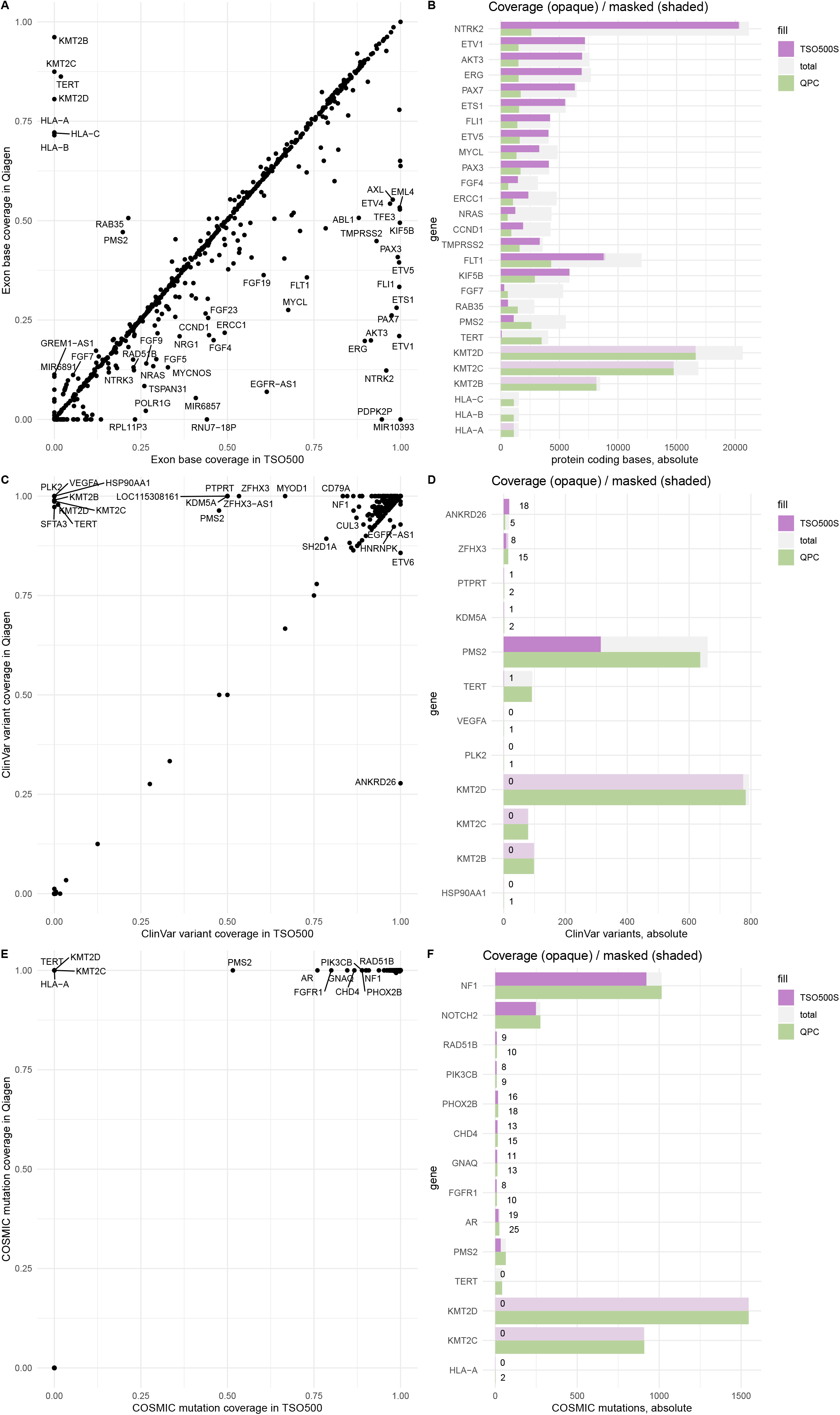
Exon base and mutation coverage in Qiagen PanCancer and TSO500 panels. Coverage of exon bases (A,B), ClinVar variants (C,D) and COSMIC CMC mutations (E,F). Note that the TSO uses a mask file to filter variant calls at positions with high error rates (shaded purple bars), but the QPC does not. Figures B, D, and F show only genes with large relative differences (filtered by fold changes of 2, 1.5, and 1.1, respectively)

ClinVar lists approximately 50,000 known variants labelled “pathogenic” or “likely pathogenic” in the advertized TSO and QPC target genes. While the TSO targeted 92.5% of these variants (94.8% without variant masking), the QPC targeted 97.4%, despite lower exon coverage. Consequently, targeting of all pathogenic variants was achieved for 182 genes in the TSO panel (200 without masking), and 223 genes in the QPC panel. In the TSO panel, no pathogenic variants were targeted in 8 genes (of which 6 were due to variant masking), whereas in the QPC it was only three. In both panels, the majority (QPC: 51%, TSO: 64%) of non-targeted variants occurred in two similar sets of only 10 genes, in both cases including NF1 and the DNA repair genes MLH1, MSH2, BRCA1, BRCA2 and ATM (Supplemental Table 1). Most differences between panels were attributable to either greater coverage in the QPC (PMS2, TERT) or extensive masking in the TSO (KMT2B, KMT2C, KMT2D) (Fig 2C, 2D).

The COSMIC cancer mutation census database (CMC, v98) lists ∼43.000 unique mutations occurring in genes targeted by the TSO and QPC panels, of which the majority (93.4%, or 99.5% without variant masking) are targeted by the TSO, and all by the QPC (100%) (Fig. 2E, 2F). Due to masking, no mutations are targeted by the TSO500 in HLA-A, KMT2C and KMT2D. Independent of masking, the QPC panel more extensively targeted mutations in NF1, TERT and PMS2 than the TSO. We estimated the frequency of samples to harbor non-targeted mutations by calculating the positive sample proportion of unique mutations in the CMC dataset, and cumulating the frequency all non-targeted mutations. The rate of non-covered CMC tier 1-3 mutations in targeted genes per sample was lower in the QPC (0.005) than in the TSO (0.14, 0.02 without masking), suggesting that one in eight (TSO, or one in 50 without masking) or one in 200 (QPC) samples would harbor oncogenic mutations that cannot be detected with the panels, in one of the panel target genes.

## Discussion

Detailed knowledge of the target regions of NGS panels is important for the correct choice and interpretation of molecular tests, but is not typically well illustrated by the test manufacturer, and usually requires bioinformatic analysis to acquire. In this study, we present PanelCat, a novel open-source tool that can be used by laboratories or NGS panel distributors to analyze NGS target regions and share this information to enable more informed decisions. A limitation of PanelCat is that it does not assess fusion or copy number events detected by panels, but respective features may be implemented in future.

PanelCat enables rapid assessment and rich visualisation of the designed target regions of NGS panels without bioinformatic expertise. As an example, we expanded in detail on the existing and incomplete documentation of two large NGS tests (Illumina TSO500 and Qiagen Pan-Cancer). PanelCat quantified precise exon coverage and identified of genes with poor coverage, extensive variant masking, differences between panels, and even discrepancies to the advertised gene list. Thus, we found that unlike the QPC, the TSO500 does not target HLA-B and HLA-C; that KMT2B, KMT2C and KMT2D are extensively masked in the TSO500 and will not yield variant calls after filtering, and that PMS2 and TERT are substantially better covered in the QPC panel independent of variant masking. In addition, we used PanelCat to describe the different coverage of individual exons of PMS2 in the panels.

PanelCat is different from the Panel Informativity Optimizer (PIO) method, previously demonstrated to assist in optimizing NGS panel design^13^. Because PanelCat does not generate new target regions, it only indirectly assists in panel design by highlighting deficits in exon or mutation coverage in particular genes of interest. Compared to PIO, PanelCat provides superior functions to analyze and compare existing (or proposed) panels. Crucially, PIO cannot process complex target regions or conventional BED-format files, only lists of complete genes or exons. Most panels target incomplete genes or exons, and would thus be inaccurately represented using PIO. Consequently, the limited panel benchmarking functions of PIO cannot inform on precise exons coverage, whereas PanelCat provides detailed information on the level of genes, exons and individual mutations. In contrast to PIO, which provides a linear data pipeline from input to output, PanelCat is a platform to collect panel analyses and visualize them for frequent inspection in a routine clinical setting. In summary, PanelCat provides opportunities that have not yet been demonstrated with previous methods, albeit with features designed more for panel end-users than panel developers.

In contrast to PIO, PanelCat does not use a variety of mutation databases to account for the heterogeneity of mutation frequencies across different disease entities. Although panels are often designed for specific disease entities or groups thereof, many widely used panels (e.g., Thermo Fisher Oncomine Focus, or Illumina TSO500) were designed to cover a wide range of disease entities, and we therefore also chose a disease-agnostic approach for PanelCat. However, the variant databases used by PanelCat can be pre-processed by users to focus the analysis entirely on mutations that are relevant in a specific disease context.

PanelCat was designed for users with limited or no IT support. For this reason, the software is designed to function on a local device (without installation of software besides R statistics); however, a slightly modified script can be hosted using ShinyServer to provide a network service. PanelCat reference databases can be easily updated, and stored panel analyses can be managed within the operating system’s file system.

Although multi-gene NGS panels are currently the standard procedure in many institutions, routine whole exome sequencing of tumor specimens is being increasingly performed. Due to the high performance of the underlying packages^5^, PanelCat could be used to analyze target regions of whole exome panels. However, the increased rendering time of some of the implemented visualization methods could be impractical. Nevertheless, the PanelCat output data, saved as R objects, could be used outside of PanelCat to plot custom graphs demanding less computation.

In conclusion, we present PanelCat as a powerful solution to current shortcomings in the presentation, analysis and awareness of NGS panel target regions. We believe this software will improve the transparency of NGS panels and facilitate more informed decisions in test choice and interpretation, thus constituting a valuable addition to the expanding repertoire of available tools.

## Data Availability

All data produced are available online at [GitHub]

## References

1. Mosele, F. et al. Recommendations for the use of next-generation sequencing (NGS) for patients with metastatic cancers: a report from the ESMO Precision Medicine Working Group. Ann. Oncol. 31, 1491–1505 (2020).

2. Niu, J., Denisko, D. & Hoffman, M. M. The Browser Extensible Data (BED) format.

3. O’Leary, N. A. et al. Reference sequence (RefSeq) database at NCBI: current status, taxonomic expansion, and functional annotation. Nucleic Acids Res. 44, D733–745 (2016).

4. R Core Team. sR: A Language and Environment for Statistical Computing. (R Foundation for Statistical Computing, 2020).

5. Lawrence, M. et al. Software for Computing and Annotating Genomic Ranges. PLoS Comput. Biol. 9, p(2013).

6. Wickham, H. ggplot2: Elegant Graphics for Data Analysis. (Springer-Verlag New York,2016).

7. Sievert, C. Interactive Web-Based Data Visualization with R, plotly, and shiny. (Chapman and Hall/CRC, 2020).

8. Chang, W. et al. shiny: Web Application Framework for R. (2022).

9. Landrum, M. J. et al. ClinVar: improvements to accessing data. Nucleic Acids Res. 48, D835–D844 (2020).

10. Tate, J. G. et al. COSMIC: the Catalogue Of Somatic Mutations In Cancer. Nucleic Acids Res. 47, D941–D947 (2019).

11. Hazini, A., Fisher, K. & Seymour, L. Deregulation of HLA-I in cancer and its central importance for immunotherapy. J. Immunother. Cancer 9, e002899 (2021).

12. Fangazio, M. et al. Genetic mechanisms of HLA-I loss and immune escape in diffuse large B cell lymphoma. Proc. Natl. Acad. Sci. 118, e2104504118 (2021).

13. Alcazer, V. & Sujobert, P. Panel Informativity Optimizer: An R Package to Improve Cancer Next-Generation Sequencing Panel Informativity. J. Mol. Diagn. 24, 697–709 (2022).

